# Victims of human trafficking and exploitation in the healthcare system: a retrospective study using a large multi-state dataset and ICD-10 codes

**DOI:** 10.1101/2022.05.02.22274579

**Authors:** Alexander Gutfraind, Kezban Yagci Sokat, Guido Muscioni, Sami Alahmadi, Jonathan Hudlow, Ronald Hershow, Beau Norgeot

## Abstract

**Background:** Trafficking and exploitation for sex or labor affects millions of persons worldwide. To improve healthcare for these patients, in late 2018 new ICD-10 medical diagnosis codes were implemented in the US.

**Methods:** Here we report on a database search of a large US health insurer that contained approximately 47.1 million patients and 0.9 million provider organizations, not limited to large medical systems. We reported on any diagnosis with the new codes between 2018-09-01 and 2022-09-01.

**Results:** The dataset was found to contain 5,262 instances of the ICD-10 codes. Regression analysis of the codes found a 5.8% increase in the uptake of these codes per year, representing a decline relative to 6.7% annual increase in the data. The codes were used by 1,810 different providers (0.19% of total) for 2,793 patients. Of the patients, 1,248 were recently trafficked, while the remainder had a personal history of exploitation. Of the recent cases, 86% experienced sexual exploitation, 14% labor exploitation and 0.8% both types. These patients were predominantly female (83%) with a median age of 20 (interquartile range: 15-35). The patients were characterized by persistently high prevalence of mental health conditions (including anxiety - 21%, post-traumatic stress disorder - 20%, major depression - 18%), sexually-transmitted infections, and high utilization of the emergency department (ED). The first report of trafficking occurred most often outside of a hospital or emergency setting (55%), primarily during primary care and psychiatric visits.

**Conclusions:** Our study strengthens the case for expanding the use of the new ICD-10 codes and studying the barriers to their implementation.

## Introduction

The United Nations defines human trafficking (also called ‘trafficking in persons’) as the “recruitment, transportation, transfer, harbouring or receipt of persons, by means of the threat or use of force … for the purpose of exploitation”(United Nations Convention against Transnational Organized Crime, 2000) and there are estimated to be tens of millions of victims worldwide (International Labor Organization, 2017; US Department of State, 2021). In the US, the National Human Trafficking Hotline reported 64,718 victims with high confidence for 2007-2020 (2020 National Hotline Annual Report, 2022), and an estimated 199,000 incidents of sexual exploitation of minors may occur each year (Estes and Weiner, 2001; Clawson et al., 2009).

The vast array of physical, sexual, psychological and social problems faced by survivors of human trafficking make it an important public health concern (Ottisova et al., 2016; Gallo et al., 2020) and healthcare providers are often on the operational frontlines when interacting with patients who are being trafficked (Ross et al., 2015; Viergever et al., 2015). However, even in clinical contexts, these patients may not make their experience known for reasons that include language or cultural barriers, fear of criminal repercussions, fear of the trafficker, or distrust of the healthcare provider (Greenbaum, 2017; Tiller and Reynolds, 2020), limiting our ability to understand their characteristics and medical needs. One emerging source of data on trafficking are a set of diagnostic ICD-10 codes introduced in late 2018. The codes are intended to be used in medical records of suspected and confirmed cases of trafficking or identify patients with a history of trafficking (Greenbaum and Stoklosa, 2019). The codes are expected to result in better coordination of care for this vulnerable population and enable new research (Macias-Konstantopoulos, 2018; Landman and Gibbs, 2021). While the new codes have been launched in the United States only, there have already been calls to expand their usage globally (Greenbaum and Stoklosa, 2019). As healthcare providers are one of the major groups caring for victims of exploitation, the new codes have the potential to provide an important new source of data about exploitation resulting from trafficking (Landman and Gibbs, 2021), and improve health outcomes for this group of patients.

Here, we aimed to characterize the uptake of the new ICD-10 codes using a comprehensive national dataset, determine in which care settings a diagnosis is typically established, and to describe the medical needs of these patients. Our literature review with Pubmed, Scopus, Web of Science and Google Scholar shows that there have been few studies based on the new ICD-10 codes. The two largest previous studies utilizing these codes relied on 49 pediatric tertiary centers (Garg et al., 2022) and 48 healthcare organizations (Kerr and Bryant, 2022). Another study (Katsanis et al., 2019) interviewed providers and examined their awareness of human trafficking but did not quantify the characteristics of the trafficked population. Additionally, a small study (n=23) reported on how the codes were used to better identify sexually exploited youth and young adults in Minnesota (Zhu et al., 2020). Our study is much larger in size (0.9 million medical organizations and four years of data) and based on a dataset that includes all providers that claimed payment for their services, regardless of size or location. It captures many more care settings including smaller providers and providers located outside major urban medical systems. Therefore, we hypothesized that the population of trafficked patients we will see will be more representative of the overall trafficked population.

## Methods

The study is a retrospective, descriptive analysis drawing from Elevance Health Inc.’s claims database. Elevance Health is one of the largest health benefits companies in the United States, managing healthcare for 47.1 million patients. The Elevance population is broadly representative of the overall US population and includes beneficiaries of commercial or government-provided health insurance across the US. The data consists of administrative claims, that is, medical bills created by medical facilities, professionals, and pharmacies and submitted to payers. In the US, the format of claims data is standardized by law (HIPAA) and contains a listing of diagnoses, health services and medications. The data includes care settings such as large medical systems, independent medical groups and individual providers, with 0.9 million provider organizations overall.

Elevance’s institutional review and ethics board deemed the study a retrospective analysis for healthcare operations in accordance with HIPAA and 45 CFR 46.102(f) and waived the requirement for patient informed consent (RITM13415607). All data utilized were de-identified and aggregated.

We performed retrospective longitudinal database search and reported on any ICD-10 diagnosis of exploitation between 2018-09-01, one month before the official launch, and 2022-09-01. After finding the claims containing the codes, we counted all patients receiving the codes. To characterize recent patients of exploitation, in the analysis we excluded patients who only had codes associated with exploitation in the past or were victims of abuse: Z62.813 / Z91.42 (personal history of exploitation) and Y07.6 (multiple perpetrators of abuse). We then compiled the medical claims for each of these patients in the 12 months before and after the trafficking event, but not within 7 days of it (to avoid inadvertently including misdated care related to the trafficking event) and reported any diagnosis and procedures codes in these intervals.

Trend was identified using regression for the daily number of bills containing the new ICD-10 codes, and separately (to avoid collinearity) regression for the number of all bills received each day. Because each medical provider has a small independent probability of using the codes on a bill, a generalized linear regression model was used. To ensure that the trend analysis is not affected by the quick uptake at the introduction of the ICD-10 codes in late 2018, the model was fit to data from 2019-01-01 through 2022-09-01. Goodness of fit testing evaluated the Negative Binomial (NB), Poisson and Gaussian models, and linear and quadratic trends (NB with a linear trend was strongly preferred).

We accessed an anonymized copy of the Elevance claims repository using a SQL client using Python 3.7, *pandas* (Reback et al., 2021) and the *sqlalchemy* package (Bayer, 2012). Regression was fitted using *statmodels* (Seabold and Perktold, 2010). For the study, the research group was provisioned with a limited view of the repository, with minimized access to clinical data. Birth dates were randomly obfuscated within a calendar year. All data was encrypted at rest and during transmission and accessed from secured devices.

## Results

The dataset contained 5,262 instances of the ICD-10 codes (Table 1).

**Table 1:** Frequency of use of the new exploitation and trafficking codes from 2018-09-01 through 2022-09-01. Total patients: 2,793. Patients with a probable recent event: 1,248 (i.e. based on any codes other than codes Y07.6, Z62.813 and Z91.42). Count gives unique bills.

| ICD-10 code | First event | Count | #Patients | Description |
| --- | --- | --- | --- | --- |
| T74.51 | 10/5/18 | 494 | 184 | Adult forced sexual exploitation, confirmed |
| T74.52 | 10/11/18 | 562 | 154 | Child sexual exploitation, confirmed |
| T74.61 | 12/7/18 | 60 | 42 | Adult forced labor exploitation, confirmed |
| T74.62 | 2/28/19 | 20 | 3 | Child forced labor exploitation, confirmed |
| T76.51 | 10/3/18 | 240 | 124 | Adult forced sexual exploitation, suspected |
| T76.52 | 10/9/18 | 321 | 151 | Child sexual exploitation, suspected |
| T76.61 | 6/29/19 | 15 | 14 | Adult forced labor exploitation, suspected |
| T76.62 | 10/13/18 | 24 | 10 | Child forced labor exploitation, suspected |
| Y07.6 | 10/3/18 | 1062 | 895 | Multiple perpetrators of maltreatment and neglect |
| Z04.81 | 10/3/18 | 759 | 520 | Encounter for examination and observation of victim following forced sexual exploitation |
| Z04.82 | 10/7/18 | 164 | 102 | Encounter for examination and observation of victim following forced labor exploitation |
| Z62.813 | 10/4/18 | 709 | 387 | Personal history of forced labor or sexual exploitation in childhood |
| Z91.42 | 10/9/18 | 832 | 372 | Personal history of forced labor or sexual exploitation |

Regression analysis found a 5.8% (p<0.05) increase in the adoption of these codes per year, representing a decline relative to the growth of the data (6.7% per year) (Figure 1).

**Figure 1:**
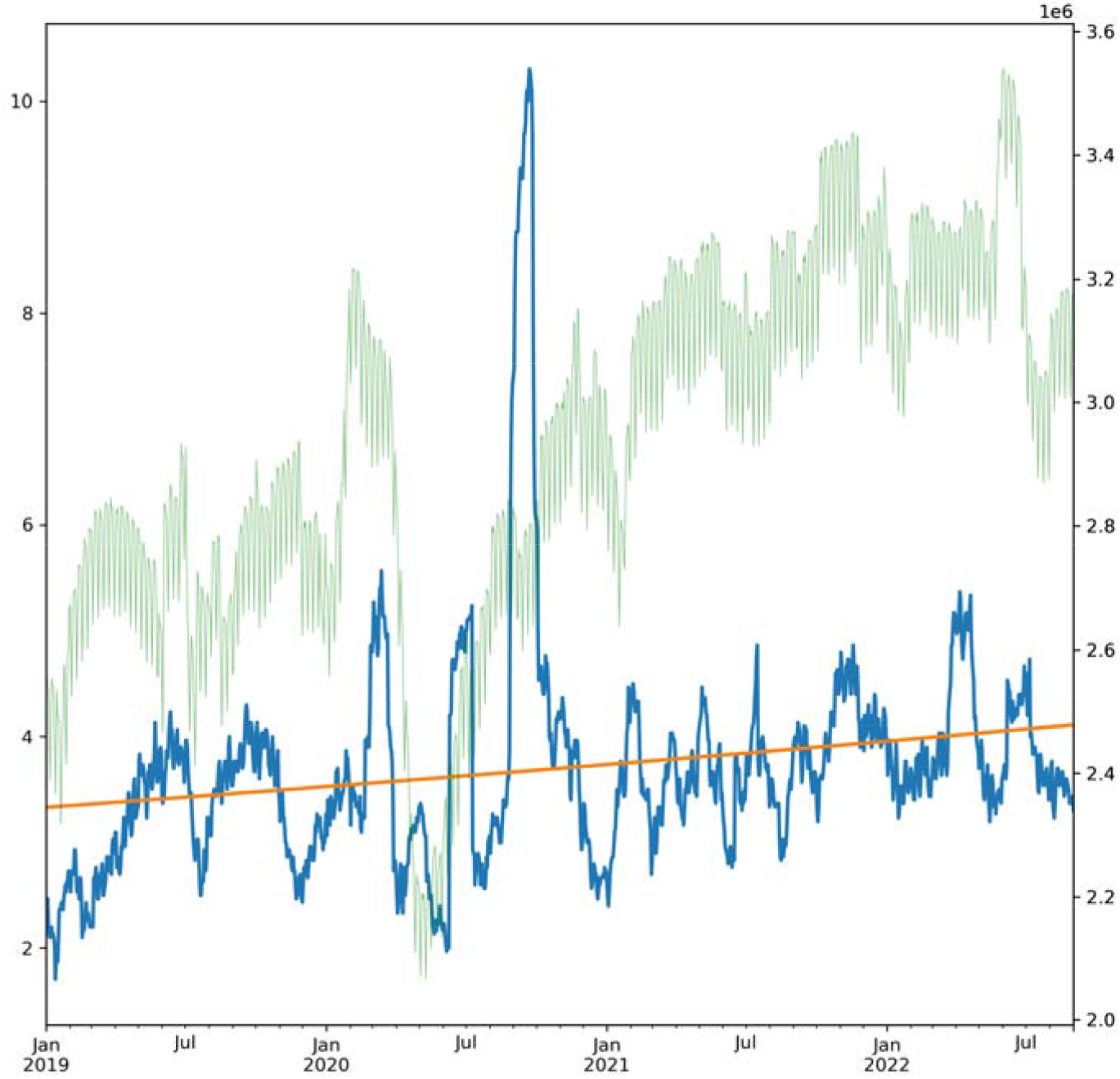
Daily count of exploitation-related ICD-10 codes from 2018-09-01 through 2022-09-01. (blue, left axis, 30-day moving average). Use of the codes increased 5.8%, representing a decrease of -0.9% per year relative to the growth of the data (6.7% per year). Superimposed trendline (orange) and overall new claims in the database (faint green, 30-day moving average).

The codes were used by 1,810 different medical providers (0.19% of total), with 77% reporting just a single patient. In 28% of the cases the codes were used as the principal or admitting diagnosis. The data contained 2,793 patients of which 1,248 were recently trafficked (reported on Table 2), while the remainder (55%) had a personal history of exploitation. Of the recently trafficked, 86% and 14% experienced sexual and labor exploitation, respectively, and 0.8% both types.

**Table 2:** Demographics of the population with new diagnoses of exploitation or trafficking. (n=1025 patients). IQR, Interquartile range.

|  | Typical value | Distribution information |
| --- | --- | --- |
| Exploitation type | 86% sexual | 14% labor, 0.8% both types of exploitation |
| Age | 20 years (median) | IQR: 15-35. Median sexual: 19.9, labor: 28.3<br>21% children under 15 and 52% under 25 |
| Gender | 82% Female | Fraction Female: Sexual: 83%, Labor: 8% |
| Race | 60% Unknown | White: 49% White, 35% Black, 11% Latin-American and 3% Asian-American |
| Health insurance | 63% Medicaid | 18% large employers, 12% multi-state employers, and others (individual policies 0.4%) |
|  | 75% Primary subscriber | 19% as child, 6% as spouse, 4% unknown/other (numbers do not sum to 100% due to rounding) |

**Table 3.** Diagnosis codes among victims of exploitation. (A) The most common diagnoses in encounters in which exploitation was first diagnosed, (B) The most common diagnoses over the preceding 12 months before the first exploitation diagnosis code, and their frequency 12 months before and after. % Pts: percent of patients with the diagnosis over the given period. Total patients: 1,248.

| (A) At establishment |  | (B) 12 month medical history |  |
| --- | --- | --- | --- |
| Description | % Pts | Description | % Pts: 12 mo before / 12 mo after |
| Encounter for examination and observation of victim following forced sexual exploitation | 42 | Encounter for immunization | 29 / 27 |
| Child sexual exploitation, suspected, initial encounter | 11 | Anxiety disorder, unspecified | 21 / 21 |
| Adult forced sexual exploitation, confirmed, initial encounter | 11 | Other long term (current) drug therapy | 21 / 20 |
| Child sexual exploitation, confirmed, initial encounter | 9 | Major depressive disorder, single episode, unspecified | 19 / 18 |
| Adult forced sexual exploitation, suspected, initial encounter | 9 | Encounter for screening for sexually transmitted infection | 17 / 15 |
| Encounter for examination and observation of victim following forced labor exploitation | 8 | Post-traumatic stress disorder, unspecified | 15 / 20 |
| Post-traumatic stress disorder, unspecified | 7 | Unspecified abdominal pain | 15 / 15 |
| Encounter for immunization | 6 | Suicidal ideations | 15 / 12 |
| Anxiety disorder, unspecified | 6 | Encounter for routine child health examination without abnormal findings | 15 / 14 |
| Encounter for screening for sexually transmitted infection | 6 | Cough | 15 / 10 |
| Suicidal ideations | 5 | Urinary tract infection, site not specified | 14 / 11 |

The patients were predominantly female (83%), insured by Medicaid (63%) and the median age was 20 (interquartile range: 15-35) with 21% children under 15 and 52% under 25. Race or ethnicity (as voluntarily self-reported during insurance enrollment) was 49% White, 35% Black, 11% Latin-American and 3% Asian-American.

Comparing the medical needs in the 12 months before and after the report of exploitation (Table 3 and Supplemental Table 1) found that the patients were characterized by persistently high prevalence of mental health conditions (including anxiety - 21%, post-traumatic stress disorder - 20%, major depression - 18%), sexually-transmitted infections, and high utilization of the emergency department (ED). Annual medical costs were high (mean $31,055 in the 12 months after diagnosis, standard deviation $152,442) compared to enrollees in Medicaid (mean $6,556 per year in 2019 - the last year with data) (Kaiser Family Foundation, 2019). The median cost was $5,254, suggesting that the costs were strongly skewed.

The first report of trafficking occurred most often outside of a hospital or emergency setting (55%): 25% during an office visit as established patients, 8% as new patients, 10% in psychiatric encounters, 4% in behavioral therapy, and the rest were diagnosed in other non-hospital settings such as community clinics. EDs accounted for just 25% of first reports.

## Discussion

In a national database, new ICD-10 codes usage reveals a population that has experienced and reported trafficking that is predominantly young and female. The population is affected by severe mental health conditions at rates many times higher than the general population (5.6% in 2020) (2020 National Survey of Drug Use and Health (NSDUH) Releases, n.d.). The population was found to be slightly older and racially more White than reported in an earlier study of Garg et al. (Garg et al., 2022), likely because Garg et al. utilized data from urban pediatric tertiary medical centers.

Generally speaking, it was surprising to see so many victims in our database, which includes just persons holding commercial or government-provided insurance, as it runs counter to the view that victims would have no insurance of any kind and receive care predominantly in emergency departments. Additionally, we were surprised to see slow uptake of the codes considering the large population of victims in the country, suggesting that there are significant barriers to adoption of the codes. While the overall volume of the data in our database grew due to increase in the enrolled population and increased medical utilization, growth in the usage of the codes was nearly 1% slower than the growth in the volume of data.

Although the patients were often treated at emergency departments, in a break with earlier studies, the majority of the patients in our data were first diagnosed outside of hospitals. Often the first diagnosis occurred as part of a routine visit as an established patient, suggesting that trust in patient-physician relationship may enable identification and reporting (Lefevre et al., 2017). The patients exhibited little or no change in their diagnoses when comparing 12 months before and after the diagnosis of exploitation, suggesting that they are unable to receive definitive care for their conditions or have chronic hard-to-treat conditions.

The available data, though large in scale, captures only a fraction of the trafficked patients nationally, and might disproportionately omit certain subgroups of patients. Underreporting might occur because patients might have barriers to accessing care, be concerned about shame, stigma, discrimination by staff or legal ramifications (Greenbaum et al., 2021). Providers might lack training on the new codes or deliberately avoid their use for the fear of potential unintended consequences, patient privacy or wrongly labeling a patient. Providers might be more likely to suspect trafficking when the patient matches a certain profile, causing underreporting of patients that fall outside that profile.

Additionally, patients and providers might be concerned about whether the trafficker has access to the medical records of the patient or not, which might make the providers hesitant about using the new ICD-10 codes (Garg et al., 2022). Because administrative claims data is used for billing, providers might sometimes be using more conventional diagnostic codes instead of trafficking, although they are incentivized to report as many medical conditions as possible. But we hypothesize that the lack of awareness and absence of institutional policies are the primary causes of underreporting, and therefore underreporting is highest in providers that see few trafficked patients. As a result, patients treated at emergency departments (EDs), youth medical centers and similar facilities are likely overrepresented in the data.

Another limitation is that the data includes only patients with medical insurance rather than those paying for care directly or receiving charitable care. The data does include patients with all types of health coverage, including employer-provided insurance, commercially-administered Medicaid and Medicare programs and individual policies. Because the health insurer is involved with patients over time and across providers, the data provides a comprehensive picture of the patients and their journeys over time. Patients without insurance likely have medical needs similar to our population but utilize medical services differently and might use emergency departments (EDs) more frequently than patients in our database. These populations are better captured in studies that rely on electronic medical record systems (Garg et al., 2022; Kerr and Bryant, 2022). COVID-19 and measures taken in some states might have impacted the uptake of codes in those states (Greenbaum et al., 2020), particularly, California and New York, which experienced lengthy shutdowns during part of the study period.

## Conclusions

Overall, our findings strengthen the case for increased vigilance for victims of trafficking in healthcare settings outside of hospitals. We agree with authorities (National Research Council and Institute of Medicine, 2014; Greenbaum et al., 2015; Shandro et al., 2016) that recommended healthcare providers should receive targeted training in safe reporting of and caring for these patients. Such interventions could increase the uptake of the codes and improve care for the patients. Our study also strengthens the case for expanding the use of the new ICD-10 codes in the US and internationally as proposed by others (Greenbaum and Stoklosa, 2019), and studying the barriers to their implementation.

## Data Availability

The data are not publicly released due to HIPAA regulations and patient privacy. The corresponding author may be contacted to discuss data access following approved agreements.

## Author Contributions

AG led the design and implementation working closely with KYS who conceived the study. GM contributed to the implementation. BN, RH, SA and JH critically reviewed the manuscript and added important intellectual content. All authors contributed to the analysis and writing and agreed to be accountable for all aspects of the work.

## Acknowledgments

AG would like to thank Stacey Sweeney for help with the data anonymization.

## References

2020 National Hotline Annual Report (2022). National Human Trafficking Hotline. Available at: https://humantraffickinghotline.org/resources/2020-national-hotline-annual-report [Accessed February 28, 2022].

2020 National Survey of Drug Use and Health (NSDUH) Releases (n.d.). Available at: https://www.samhsa.gov/data/release/2020-national-survey-drug-use-and-health-nsduh-releases [Accessed December 21, 2022].

Bayer, M. (2012). “SQLAlchemy,” in The Architecture of Open Source Applications Volume II: Structure, Scale, and a Few More Fearless Hacks, eds. A. Brown and G. Wilson (aosabook.org).

Clawson, H. J., Dutch, N., Solomon, A., and Grace, A. L. G. (2009). Human Trafficking Into and Within the United States: A Review of the Literature. ASPE. Available at: https://aspe.hhs.gov/reports/human-trafficking-within-united-states-review-literature-0 [Accessed February 28, 2022].

Estes, R. J., and Weiner, N. A. (2001). The Commercial Sexual Exploitation of Children in the U.S., Canada and Mexico: Final Report (of the U.S. National Study).

Gallo, M., Konrad, R. A., and Thinyane, H. (2020). An Epidemiological Perspective on Labor Trafficking. Journal of Human Trafficking, 1–22.

Garg, A., Panda, P., Malay, S., and Slain, K. N. (2022). Human trafficking ICD-10 code utilization in pediatric tertiary care centers within the United States. Front. Pediatr. 10. doi: 10.3389/fped.2022.818043.

Greenbaum, J., Crawford-Jakubiak, J. E., and Committee on Child Abuse and Neglect (2015). Child sex trafficking and commercial sexual exploitation: health care needs of victims. Pediatrics 135, 566–574.

Greenbaum, J., McClure, R. C., Stare, S., Barnes, W., Claire E. Castles, J. D., Culliton, E. R., et al. (2021). Documenting ICD Codes and Other Sensitive Information in Electronic Health Records. Available at: https://www.mhaonline.org/docs/default-source/resources/human-trafficking/icd-codes-and-other-sensitive-information-in-ehr.pdf [Accessed April 27, 2021].

Greenbaum, J., and Stoklosa, H. (2019). The healthcare response to human trafficking: A need for globally harmonized ICD codes. PLoS Med. 16, e1002799.

Greenbaum, J., Stoklosa, H., and Murphy, L. (2020). The Public Health Impact of Coronavirus Disease on Human Trafficking. Front Public Health 8, 561184.

Greenbaum, V. J. (2017). Child sex trafficking in the United States: Challenges for the healthcare provider. PLoS Med. 14, e1002439.

International Labor Organization (2017). Statistics on forced labour, modern slavery and human trafficking (Forced labour, modern slavery and human trafficking) in 2016. Available at: http://www.ilo.org/global/topics/forced-labour/statistics/lang--en/index.htm [Accessed February 28, 2022].

Kaiser Family Foundation (2019). State Health Facts - Medicaid spending per enrollee. KFF. Available at: https://www.kff.org/medicaid/state-indicator/medicaid-spending-per-enrollee/ [Accessed March 7, 2022].

Katsanis, S. H., Huang, E., Young, A., Grant, V., Warner, E., Larson, S., et al. (2019). Caring for trafficked and unidentified patients in the EHR shadows: Shining a light by sharing the data. PLoS One 14, e0213766.

Kerr, P. L., and Bryant, G. (2022). Use of ICD-10 Codes for Human Trafficking: Analysis of Data From a Large, Multisite Clinical Database in the United States. Public Health Rep. 137, 83S–90S.

Landman, A., and Gibbs, H. (2021). ICD Codes - An Important Component for Improving Care and Research for Patients Impacted by Human Trafficking. J. Law Med. Ethics 49, 290–292.

Lefevre, M., Hickle, K., Luckock, B., and Ruch, G. (2017). Building trust with children and young people at risk of child sexual exploitation: The professional challenge. Br. J. Soc. Work 47, 2456–2473.

Macias-Konstantopoulos, W. L. (2018). Diagnosis Codes for Human Trafficking Can Help Assess Incidence, Risk Factors, and Comorbid Illness and Injury. AMA J Ethics 20, E1143–1151.

National Research Council, and Institute of Medicine (2014). Confronting Commercial Sexual Exploitation and Sex Trafficking of Minors in the United States: A Guide for the Health Care Sector. National Academies Press.

Ottisova, L., Hemmings, S., Howard, L. M., Zimmerman, C., and Oram, S. (2016). Prevalence and risk of violence and the mental, physical and sexual health problems associated with human trafficking: an updated systematic review. Epidemiol. Psychiatr. Sci. 25, 317–341.

Reback, J., McKinney, W., Van den Bossche, J., Augspurger, T., Cloud, P., Hawkins, S., et al. (2021). pandas-dev/pandas: Pandas 1.3.3. Zenodo doi: 10.5281/ZENODO.3509134.

Ross, C., Dimitrova, S., Howard, L. M., Dewey, M., Zimmerman, C., and Oram, S. (2015). Human trafficking and health: a cross-sectional survey of NHS professionals’ contact with victims of human trafficking. BMJ Open 5, e008682.

Seabold, S., and Perktold, J. (2010). Statsmodels: Econometric and statistical modeling with python. in Proceedings of the 9th Python in Science Conference (SciPy). doi: 10.25080/majora-92bf1922-011.

Shandro, J., Chisolm-Straker, M., Duber, H. C., Findlay, S. L., Munoz, J., Schmitz, G., et al. (2016). Human Trafficking: A Guide to Identification and Approach for the Emergency Physician. Ann. Emerg. Med. 68, 501–508.e1.

Tiller, J., and Reynolds, S. (2020). Human Trafficking in the Emergency Department: Improving Our Response to a Vulnerable Population. West. J. Emerg. Med. 21, 549–554.

United Nations Convention against Transnational Organized Crime (2000). United Nations : Office on Drugs and Crime. Available at: https://www.unodc.org/unodc/en/organized-crime/intro/UNTOC.html [Accessed February 28, 2022].

US Department of State (2021). Trafficking in Persons Report. Available at: https://www.state.gov/wp-content/uploads/2021/09/TIPR-GPA-upload-07222021.pdf.

Viergever, R. F., West, H., Borland, R., and Zimmerman, C. (2015). Health care providers and human trafficking: what do they know, what do they need to know? Findings from the middle East, the Caribbean, and central america. Front Public Health 3, 6.

Zhu, L., Roesler, J., Menanteau, B., and Kindemark, M. (2020). 87 The use of hospital discharge data to count sexually exploited youth and young adults in Minnesota. in Poster Presentations (BMJ Publishing Group Ltd). doi: 10.1136/injuryprev-2020-savir.106.

